# Sex-specific epidemiological and clinical characteristics of Covid-19 patients in the southeast region of Bangladesh

**DOI:** 10.1101/2021.07.05.21259933

**Authors:** Md. Aminul Islam, Abdullah Al Marzan, Md. Sydul Islam, Samina Sultana, Md. Iftakhar Parvej, Mohammad Salim Hossain, Mohammad Tohid Amin, Farzana Ehetasum Hossain, Md Abdul Barek, Md. Shafiul Hossen, Md. Shariful Islam, Foysal Hossen, Newaz Mohammed Bahadur, Md. Shahadat Hossain, Nazratun Nayeem Choudhury, Md. Didar-ul Alam, Firoz Ahmed

## Abstract

**Background:** COVID-19 has become a global pandemic with a high growth rate of confirmed cases. In Bangladesh, both mortality and affected rates are increasing at an alarming rate.

Therefore, more comprehensive studies of the epidemiological and clinical characteristics of COVID-19 are required to control this pandemic.

**Purpose:** The present study aimed to compare and analyze the sex-specific epidemiological, clinical characteristics, comorbidities, and other information of confirmed COVID-19 patients from the southeast region in Bangladesh for the first time.

**Methods:** 385 lab-confirmed cases were studied out of 2,471 tested samples between 5 June and 10 September 2020. RT-PCR was used for COVID-19 identification and SPSS (version 25) for statistical data analysis.

**Results:** We found that male patients were roughly affected compared to females patients (male 74.30% vs. female 25.7%) with an average age of 34.86 ± 15.442 years, and B (+ve) blood group has been identified as a high-risk factor for COVID-19 infection. Workplace, local market, and bank were signified as sex-specific risk zone (*p* < 0.001). Pre-existing medical conditions such as diabetes, hypertension, cardiovascular and respiratory diseases were identified among the patients. Less than half of the confirmed COVID-19 cases in the southeast region were asymptomatic (37.73%) and more prevalent among females than males (male vs. female: 36.84% vs. 40.51%, *p* = 0.001).

**Conclusions:** The findings may help health authorities and the government take necessary steps for identification and isolation, treatment, prevention, and control of this global pandemic.

## 1 Introduction

COVID-19 (Coronavirus disease-2019) was first identified in December 2019 in a person at a seafood market in Wuhan city, in Hubei province, China (Chan et al., 2020). It is caused by an unknown pathogenic virus, and somedays later, scientists isolated this new virus from the lower respiratory tract of patients. This virus is a new type of Beta coronavirus, corresponds to a comparatively noted Coronaviridae family, and is identical to Bat SARS-type coronavirus (having 88% identity) and have identity nucleotide with the actual severe acute respiratory syndrome (SARS) (80%) and Middle East respiratory syndrome coronavirus (MERS-CoV) (about 50%) epidemic virus (Zheng, 2020). The pathogen of this disease has been confirmed by molecular methods as a new coronavirus and was primarily named a 2019 novel coronavirus (2019-nCoV). World Health Organization (WHO) declared the COVID-19 outbreak as the sixth public health emergency of international concern on 30 January 2020 and declared a fresh name, called Corona Virus Disease (COVID-19) on 11 February 2020, for the pandemic disease: Corona Virus Disease (COVID-19) (Lai et al., 2020).

On the grounds of taxonomy, phylogeny, and developed practice, the International Committee on Taxonomy of Viruses (ICTV) announced the official name of this coronavirus as severe acute respiratory syndrome coronavirus 2 (SARS-CoV-2) on 12 February 2020 (Lai et al., 2020). Since the disease’s occurrence, it is become a global threat and spreading quickly to provinces in China and more than 210 countries, including Bangladesh, worldwide (Worldometers, n.d.). By 2 January 2020, 41 lab-confirmed COVID-19 patients had been detected from hospitals, and less than 50% of them had the implicit illness, including high blood pressure, hyperglycemia, and cardiovascular diseases (C. Huang et al., 2020). As of 22 January 2020, there were 571 registered cases of 2019-new coronaviruses (COVID-19) in 25 provinces in China and 17 deaths (Lu, 2020). Now, 4636 patients died up to 22 July 2021 in China (Worldometers, n.d.).

COVID-19 infects more than 19.3 million patients, and about 4,147,24 (0.22%) died on 22 July 2021 worldwide. USA has accounted for the highest number of COVID-19 patients (35,147,918), followed by India (31,289,115), Brazil (19,474,489), Russia (6,030,240), and the France (5,911,601) on 22 July 2021. Every country in South Asia is affected by this zoonotic virus. About 31,289,115 cases in India have been identified, one of the top five affected countries (Worldometers, n.d.). Bangladesh announced the first confirmed coronavirus cases on 8 March 2020, and this number increased day by day (Anwar et al., 2020). It was reported that about 1,136,503 cases were identified, 18.498 patients died, and 961,044 patients are recovered by 22 July 2021 (Worldometers, n.d.). It is now spreading all over the country, and the highest affected area was observed in the Dhaka division. Based on district-wise case distribution, around 5546 and 2309 patients were identified, and 5058 and 2232 patients were recovered in Noakhali and Lakshmipur, respectively, on 31 December 2020 (Civil Surgeon Office, Noakhali, and Laxmipur, 2020, Corona Information, n.d.). It diffuses via people-to-people transmission, for instance, sneezing and coughing, while some research indicates the transmission chances even amongst the asymptomatic (Ningthoujam, 2020). It can also spread through stool, sweat, urine, and respiratory secretions.

The general incubation period of this virus is 1 to 14 days (Baud et al., 2020); however, a longer latent period has been reported in some cases, which makes it more dangerous as the host may transmit the disease to others unknowingly (Kong, 2020). The most frequently occurring symptoms are fever, cough, sore throat, tiredness, ache, headache, etc. Still, severe illnesses, including-diarrhea, pneumonia, pulmonary edema, acute respiratory distress syndrome, multiple organ failure, are also found among patients (Chen et al., 2020; Y. Meng et al., 2020). Both males and females are affected by COVID-19, but several epidemiological and clinical studies suggest that males are more progressive to infection than females (Barek et al., 2020; Y. Meng et al., 2020). Many studies have already found that single or multiple comorbidities like high blood pressure, cardiovascular disease, high blood pressure, liver problem, diabetes, lung disease, and kidney disease patients are more affected by this virus (Barek et al., 2020; Emami et al., 2020). Researchers are trying to develop medicine and vaccine to combat COVID-19 and upcoming SARS and MERS viruses (Amanat and Krammer, 2020). Now, public concern and consciousness are the main prohibitions for the COVID-19 outbreak.

At present, epidemiological, clinical characteristics, the pervasiveness of comorbidities, other information is most important to take necessary steps for treatment, prevention, and control of the current outbreak. There have no published demographic and clinical characteristics study data regarding COVID-19 patients of Bangladesh. Considering the above-mentioned facts, we analyzed 385 laboratory-confirmed COVID-19 positive patients’ demographic data, clinical manifestations, medical history, blood group, travel history, previous vaccination, and other data from the southeast region in Bangladesh for the very first time, which help to proper decisions to reduce the risk of infection.

## 2 Methods

### 2.1 Study design and participants

We selected only laboratory-confirmed Covid-19 cases between 5 June 2020 to 10 September 2020 who were tested at Noakhali Science and Technology University COVID-19 testing laboratory, Noakhali, Bangladesh approved by the government (The Daily Star, 2020). The specimen was collected by nasopharyngeal swab, and reverse transcriptase-polymerase chain reaction (RT-PCR) assay was used for Covid-19 confirmation. Three hundred eighty-five patients (aged above 5 years) were identified as positive from the southeast region, Bangladesh (Noakhali and Lakshmipur) in where 286 patients were male and 99 were female. Pregnant women, children, adults, and older patients were included in this study. To avoid the chance of bias, participants were selected and listed with the help of the Noakhali Science and Technology University Covid-19 testing laboratory database.

### 2.2 Data collection

The questionnaire was adopted from formerly published studies, and our research team added, modified, and developed some questions. There were various sections in the survey, including demographic information, clinical characteristics, pre-existing medical condition, blood group, earlier visited information, vaccination, etc. The investigators were told that their participation was anonymous and entirely voluntary, and there was no reward for taking part. All the participants willingly participated in this work by giving written permission. They were invited to complete the questionnaire. We were present on hand to answer questions or clarify any doubts that they might have. All filled questionnaires were collected by us one by one.

### 2.3 Definition

Patients with SARS-CoV-2 infection have no clinical signs and symptoms were defined as asymptomatic cases, and symptomatic patients were defined with SARS-CoV-2 infection presenting with clinical characteristics from medical interviews. Comorbidities were ascertained from physician documentation.

### 2.4 Specimen collection and RT-PCR assay for SARS-CoV-2

We detected the presence of SARS-CoV-2 by the qPCR method with the novel coronavirus (2019-nCoV) nucleic acid diagnostic Kit (PCR-fluorescence probing). The test utilizes the novel coronavirus (2019-nCoV) ORF 1ab and the specific conserved sequence of coding nucleocapsid protein N gene as the target regions to detect sample RNA through fluorescence signal changes. These diagnostic criteria were based on the recommendations by the Institute of Epidemiology, Disease Control and Research (IEDCR), Bangladesh. Test facility and quality reconfirmed by IEDCR.

### 2.5 Statistical Analysis

Descriptive statistics were used to summarize the data, continuous variables were expressed as mean ±SD, and categorical variables were summarized as counts and percentages. There are several kinds of models for analyzing the relationship between the data sets. As our data are parametric, we used Pearson’s Correlation Coefficient (2-tailed). All statistical analysis was conducted using the IBM Statistical Package for the Social Sciences (SPSS), version 25.0 software. P < 0.05 was considered statistically significant. Correlation between sex and many other factors i.e. BMI, age, comorbidities, etc., was analyzed and determined.

## 3 Results

The demographic characteristics of all the confirmed COVID-19 cases (N=385) were shown in **Table 1**. Of them, 74.3% of cases were male, and 25.7% were female, with an average age of 34.86 ± 15.442 years. The highest number of patients was in the age range of 21-50 years (71.91%). However, around 15% of cases were found older than them. As for Body Mass Index (BMI), 23.80% of cases were overweight, and 8.10% were obese, while only 3.40% were underweight. A (+ve), B (+ve), and O (+ve) blood groups were the most identified blood group with a percentage of 25.46%, 32.52%, and 18.40%, respectively. In the meanwhile, only a few percent of patient cases were observed with O (-ve) (3.07%), B (-ve) (3.68%), and A (-ve) (3.37%) blood group. Of the male with a positive blood group, the most common blood group was B (+ve) (35.22%), followed by A (+ve) (22.76%) and AB (+ve) (16.46%). In the female positive blood group, the most specific blood group was still B (+ve) and O (+ve) (24.05%) in each case, followed by A (+ve) (21.05%) and AB+ (15.79%) (**Figure 1**).

**Table 1:** Demographic characteristics of COVID-19 cases from the southeast region in Bangladesh.

| **Characteristics** | **Response** | **Patients** |
| --- | --- | --- |
| **Sex (N = 385)** | Male | 286 (74.3%) |
|  | Female | 99 (25.7%) |
| **Age (years) (N=381)** | Mean ± SD | 34.86 ± 15.442 |
| **Age group** | 0-20 | 50(13.12%) |
|  | 21-50 | 274 (71.91%) |
|  | >51 | 57 (14.96%) |
| **BMI (N= 298)** | Mean ± SD | 24.1012 ± 4.117 |
| **BMI range** | <18.5 (Underweight) | 10 (3.40%) |
|  | 18.5-24.9 (Normal) | 193 (64.7%) |
|  | 25-29.9 (Overweight) | 71 (23.80%) |
|  | >30 (Obese) | 24 (8.10%) |
| **Blood group (N=326)** | A (+ve) | 83(25.46%) |
|  | B (+ve) | 106 (32.52%) |
|  | AB (+ve) | 43 (13.19%) |
|  | O (+ve) | 60 (18.40%) |
|  | A (-ve) | 11 (3.37%) |
|  | B (-ve) | 12 (3.68%) |
|  | AB (-ve) | 1 (0.31%) |
|  | O (-ve) | 10 (3.07%) |
| **Educational status (N= 298)** | Masters | 35 (11.75%) |
|  | Graduate | 76 (25.50%) |
|  | Degree | 8 (2.68%) |
|  | HSC | 85 (28.50%) |
|  | SSC | 28 (9.40%) |
|  | Below SSC | 46 (15.44%) |
|  | No education | 20 (6.73%) |
| **Occupation (N=298)** | Civil servant | 42 (14.09%) |
|  | Health worker (doctor, nurse,<br>sample collector) | 24 [7,8,9] (8.05%)<br>[2.68%,2.35%,3.02%] |
|  | Engineer | 1 (0.33%) |
|  | Banker | 8 (2.68%) |
|  | Private job | 43 (14.43%) |
|  | Businessmen | 56 (18.80%) |
|  | Homemaker | 42 (14.09%) |
|  | Student | 46 (15.43%) |
|  | Others | 36 (12.08%) |
| **Traveling to other countries**<br>**(N=298)** |  |  |
|  | Yes | 6 (2.01%) |
|  | No | 292 (97.99%) |
| **Earlier visited places (N=326)** |  |  |
|  | Workplace | 215 (65.95%) |
|  | Bank | 64 (19.63%) |
|  | Local market | 148 (45.39%) |
|  | Relatives house | 59 (18.09%) |
|  | Social gathering | 36 (11.04%) |
|  | Health care center | 45 (13.80%) |
| **Comorbidities (N= 385)** |  |  |
|  | Diabetes | 20 (5.19%) |
|  | Hypertension | 23 (5.97%) |
|  | Asthma | 29(7.53%) |
|  | Allergy | 2(0.52%) |
|  | Cancer | 3(0.78%) |
|  | Pregnancy | 9 (2.34%) |
|  | Others | 5 (1.29%) |
|  | No risk factors | 296 (76.35%) |
| **Vaccine in life (N=298)** |  |  |
|  | BCG | 257 (86.2%) |
|  | Polio | 262 (87.9%) |
|  | Hepatitis B | 44 (14.8%) |
|  | Mumps | 161 (54.00%) |
|  | Tetanus | 246 (82.5%) |
|  | Chickenpox | 45 (15.1%) |
|  | Measles | 9 (3.00%) |
| **Death Case** |  |  |
| **Age (years) (N=130)** | Mean ± SD | 62.56 ± 14.252 |
| Age group | 0-20 | 1 (0.77%) |
|  | 21-50 | 23 (17.70%) |
|  | >51 | 106 (81.54%) |

**Figure 1.**
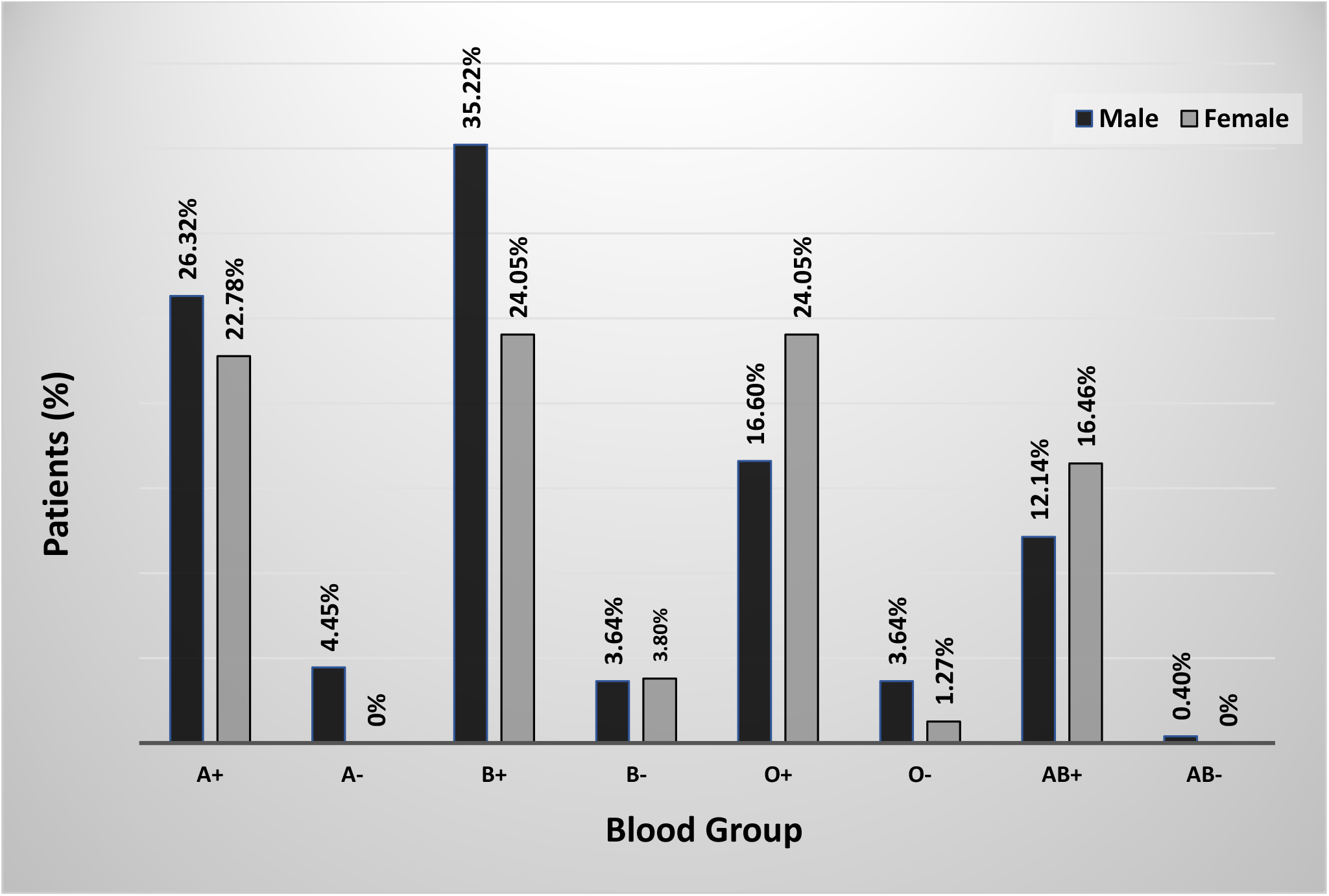
Percentage of sex-specific blood group of COVID-19 infected patients

About 28.50% of cases passed the higher secondary certificate (HSC) level, while only 9.40% continued their education until the secondary school certificate (SSC) level. Graduated (25.50%) and post graduated (11.75%) cases were also observed in our study. Notably, most of the COVID-19 patients were businessmen (18.80%), students (15.43%), civil servants (14.09%), and health workers (8.05%). In male cases, 23.04% were businessmen, followed by 17.83% civil servants and 8.26% health workers. However, two-thirds of the female cases (61.76%) were housewives, followed by 20.59% of students (**Figure 2**). All confirmed cases ensured that they did not travel to any other countries within the nearest time before being infected. However, most of them visited their workplace (65.95%), local market (45.39%), and health care center (13.80%) within the earlier time. About 19.63% and 18.09% of cases visited the bank and relative house, respectively. Our study also explored that only 15.50% of infected persons had clear contact with confirmed cases as far as they know. Additionally, diabetes (5.19%), hypertension (5.97%), asthma (7.53%), allergy (0.52%) cancer (0.78%), pregnancy (2.34%) and others (1.29%) were the most common comorbidities identified among the confirmed cases. Most of the patients were vaccinated with BCG (86.2%), Polio (87.9%), Mumps (54%), Tetanus (82.5%). However, only a few of them were vaccinated with Hepatitis B (14.8%), Chickenpox (15.1%), and Measles (3%).

**Figure 2.**
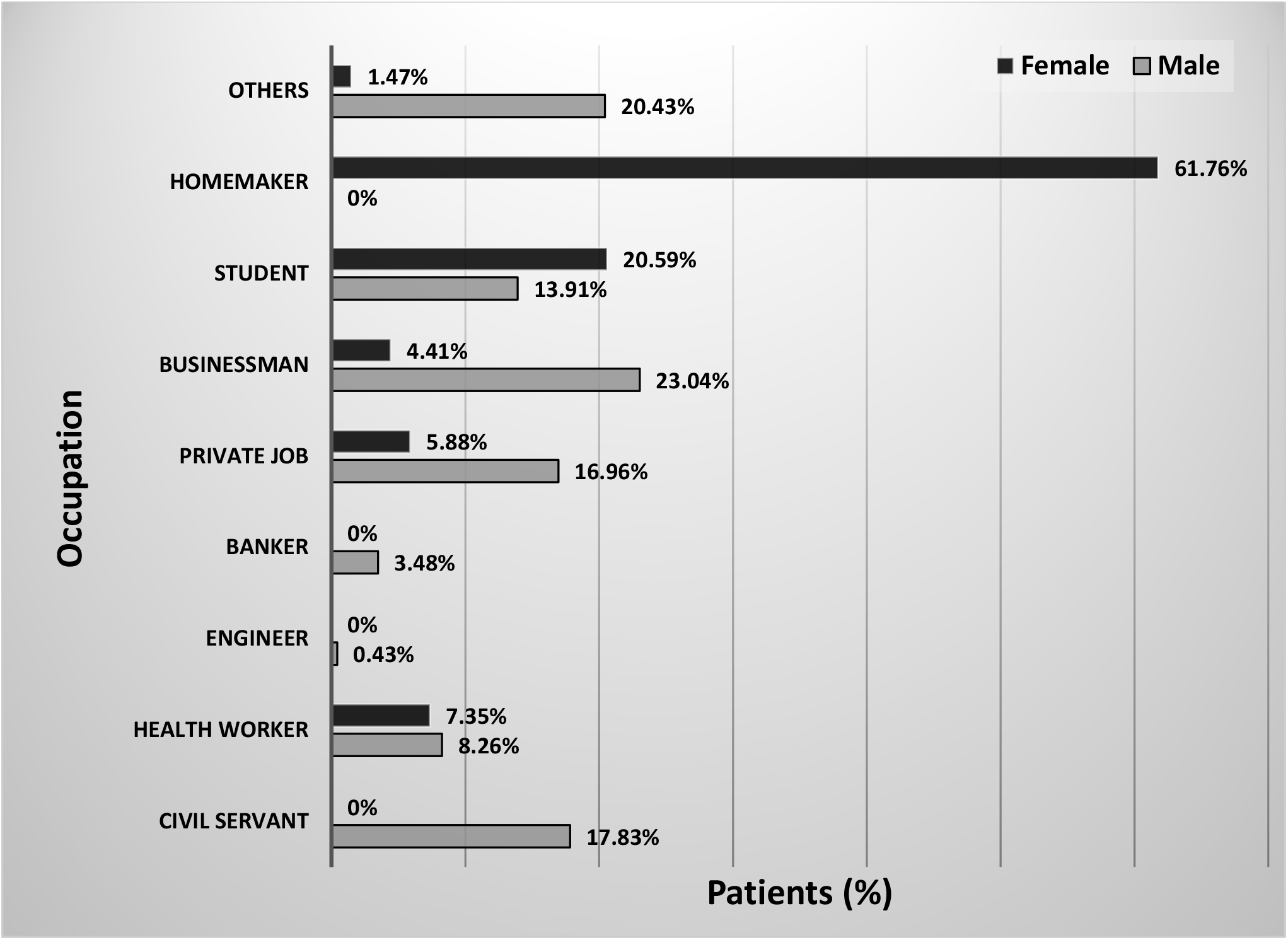
Percentage of sex-specific occupations of COVID-19 patients

Analysis of demographic characteristics based on gender-specific is explored in **Table 2**. This analysis showed that BMI and earlier visited places showed significant (*p* < 0.05) differences between male and female patients. The highest percent of both males (74.24%) and females (68.1%) were in the age range of 21-50 years. About 88.66% male cases and 92.40% female cases had positive Rh factor (*p* = 0.115). Additionally, earlier visited place such as workplace (*p* < 0.001), bank (*p* < 0.001), local market (*p* < 0.001), and social gathering (*p* < 0.001) were also reported as the significant risk factors for sex-specific COVID-19 infection. Among all comorbidities, our study found significant differences between males and females in the case of only chickenpox (*p* = 0.014).

**Table 2:** Sex-specific analysis of demographic characteristics of confirmed COVID-19 cases.

| Category | Male | Female | *p*-value |
| --- | --- | --- | --- |
| Age group (Years) (M=284, F= 97) |  |  |  |
| 0-20 | 30 (10.56%) | 20 (20.6%) | 0.275 |
| 21-50 | 208 (73.24%) | 66 (68.1%) |  |
| >51 | 46 (16.20%) | 11 (11.3%) |  |
| BMI range (M=230, F= 68) |  |  |  |
| <18.5 (Underweight) | 6 (2.61%) | 4 (5.9%) | < 0.001* |
| 18.5-24.9 (Normal) | 152 (66.09%) | 41 (60.29%) |  |
| 25-29.9 (Overweight) | 52 (22.60%) | 19 (27.94%) |  |
| >30 (Obese) | 17 (8.70%) | 4 (5.88%) |  |
| Rh factor (M=247, F=79) |  |  |  |
| Positive | 219 (88.66%) | 73 (92.40%) | 0.115 |
| Negative | 28 (11.34%) | 6 (7.60%) |  |
| Traveling to other countries<br>(M= 230, F= 68) |  |  |  |
| Yes | 5 (2.17%) | 1 (1.47%) | 0.769 |
| No | 225 (97.83%) | 67 (98.53%) |  |
| Earlier visited places<br>(M=229, F=97) |  |  |  |
| Workplace | 190 (82.97%) | 25 (25.77%) | < 0.001* |
| Bank | 55 (24.01%) | 9 (9.28%) | < 0.001* |
| Local market | 133 (58.08%) | 15 (15.46%) | < 0.001* |
| Relatives house | 38 (16.60%) | 21 (21.65%) | 0.404 |
| Social gathering | 34 (14.85%) | 2 (2.06%) | < 0.001* |
| Health care center | 29 (12.66%) | 16 (16.49%) | 0.400 |
| Comorbidities (M= 288, F=97) |  |  |  |
| Diabetes | 14 (4.86%) | 6 (6.18%) | 0.612 |
| Hypertension | 21 (7.29%) | 2 (2.06%) | 0.060 |
| Asthma | 24 (8.33%) | 5 (5.15%) | 0.306 |
| Cancer | 2 (0.01%) | 1 (1.00%) | 0.745 |
| Pregnancy | --- | 9 (9.27%) | < 0.001* |
| Others | 3 (1.04%) | 2 (2.06%) | 0.444 |
| No risk factors | 224 (77.78%) | 72 (74.22%) | --- |
| **Vaccine in life (M=230, F=68)** |  |  |  |
| BCG | 81 (100.00%) | 55 (80.9%) | 0.084 |
| Polio | 81 (100.00%) | 59 (86.8%) | 0.519 |
| Hepatitis B | 12 (14.81%) | 14 (20.6%) | 0.159 |
| Mumps | 77 (95.06%) | 40 (58.8%) | 0.551 |
| Tetanus | 75 (92.59%) | 54 (79.4%) | 0.318 |
| Chickenpox | 1 (1.23%) | 17 (25.00%) | 0.014** |
| Measles | 7 (3.04%) | 2 (2.94%) | 0.928 |
| Diphtheria | 196 (85.22%) | 55(80.9%) | 0.269 |

\* Correlation is significant at the 0.01 level (2-tailed)
\*\* Correlation is significant at the 0.05 level (2-tailed)

Signs and symptoms identified during testing among COVID-19 confirmed cases are tabulated in **Table 3**. Our study also confirmed the sex-specific clinical characteristics of COVID-19. This present study found that less than half of the confirmed COVID-19 cases (37.73%) were symptomatic, and this asymptomatic characteristic was more prevalent among females (40.51%) than that of males (36.84%) patients (*p* = 0.001). Conversely, mortality cases were found higher for males than females with no significant correlation (*p* = 0.724). Of the patients showing symptoms, the most frequent symptom was fever (29.45%), followed by cough (34.05%) and headache (13.50%), with no significant difference between genders. Fever and cough were observed mainly among females (55.32% and 44.87%) than male patients (44.87% and 53.85%). Also, 8.28% of cases and 13.50% of cases experienced sore throat and headache. Though very few female cases showed fever (*p* = 0.240); however, about 37.82% of male cases experienced sore throat in comparison to only 25.53% of female cases (*p* = 0.204).

**Table 3:** Sex-specific signs and symptoms of COVID-19 patients identified during testing.

| Items | Total (%) | Male (%) | Female (%) | p-value |
| --- | --- | --- | --- | --- |
| Asymptomatic | 123 (37.73%) | 91 (36.84%) | 32 (40.51%) | 0.001<sup>*</sup> |
| Symptomatic | 203 (62.27%) | 156 (63.16%) | 47 (59.49%) |  |
| **Symptoms** |  |  |  |  |
| **(M=156, F=47)** |  |  |  |  |
| Fever | 96 (29.45%) | 70 (44.87%) | 26 (55.32%) | 0.240 |
| Cough | 111 (34.05%) | 84 (53.85%) | 27 (57.45%) | 0.666 |
| Sore throat | 27 (8.28%) | 22 (14.10%) | 5 (10.64%) | 0.542 |
| Headache | 44 (13.50%) | 34 (21.79%) | 10 (21.27%) | 0.817 |
| Tiredness | 71 (21.78%) | 59 (37.82%) | 12 (25.53%) | 0.204 |
| Ache | 41 (12.58%) | 28 (17.95%) | 13 (27.66%) | 0.080 |
| Diarrhea | 10 (3.07%) | 7 (4.48%) | 3 (6.38%) | 0.601 |
| **Death significance** |  |  |  |  |
| **(M=108, F=22)** |  |  |  |  |
| **Age (years)** |  |  |  |  |
| 0-20 | 1 (0.77%) | 1 (0.93%) | 0 (0.00%) | 0.724 |
| 21-50 | 23 (17.70%) | 17 (15.70%) | 6 (27.27%) |  |
| >50 | 1 (0.77%) | 90 (83.33%) | 16 (72.72%) |  |

<sup>\*</sup>Correlation is significant at the 0.01 level (2-tailed)

## 4 Discussion

The new beta coronavirus is the third type of zoonotic coronavirus, the seventh human coronavirus, and has similarities with previous coronavirus (Zhou et al., 2020; Zhu et al., 2020). This contagious virus shows the same receptor binding domain with SARS coronavirus (SARS-CoV) and MERS-CoV (Lu et al., 2020). The number of COVID-19 cases continuously rising worldwide, but no specific treatment has been confirmed to be fully effective against COVID-19 (irrespective of different strains). Therefore, it is crucial to identify the clinical demographical characteristics, clinical manifestation, comorbidities, and other information of COVID-19 patients. It is also important to help in the early detection and isolation of infected individuals and minimize the spread of the disease, severity, and death rate. To our best knowledge, this is the first study undertaken to investigate the demographic, clinical characteristics, comorbidities, and excrescent information of the confirmed COVID-19 patients from the southeast region in Bangladesh.

We analyzed 200 positive samples from Noakhali and 185 positive samples from Lakshmipur, the southeast region of Bangladesh. In our study, we observed that the number of male patients was largely (74.3%), almost three times higher than female patients (25.7%). Several studies conducted in China and other countries corroborated a higher number of infected males than that of females (Novel, 2020; Z. Wang et al., 2020). Meanwhile, a research study by Zhou *et al*. suggests that the expression of ACE2, the receptor for COVID-19, is higher in males than that in females, which may be the reason behind the higher proportion of COVID-19 infected males (Zhou et al., 2020). It is also thought that as females have more X chromosomes and sex hormones, these factors play an essential role in innate and adaptive immunity (Jaillon et al., 2019). On the other hand, Females are less associated with a bad lifestyle than males. Similarly, several studies found that males are more infected than females during MERS-CoV and SARS-CoV pandemic (Badawi and Ryoo, 2016; Channappanavar et al., 2017). Additionally, the average age of all cases was 34.86 ± 15.442 years old and nearly similar to several studies (Azlan et al., 2020; Q. Huang et al., 2020; Kim et al., 2020). It might be due to the outgoing of middle-aged people for their respective work or for their family need as they have to meet all family and society demands. However, our study indicated that patients at a wide age range can be infected by SARS-CoV-2. This present study also demonstrated a slightly greater mean BMI value than those found in the Hubei (Yang et al., 2020) and Jiangsu (R. Huang et al., 2020) provinces of China. Notably, more than three fourth of patients involved in our study had a positive blood group, including A (+ve) (25.46%), B (+ve) (32.52%), and O (+ve) (18.40%), similar to Li *et al*. and Zietz *et al*. studies (Li et al., 2020; Zietz et al., 2020). According to one study, anti-A antibodies inhibit S protein/angiotensin-converting enzyme 2-dependent adhesion of these cells to an angiotensin-converting enzyme 2 expressing cell line (Guillon et al., 2008).

Besides, the majority of infected cases were business professionals (18.80%), followed by students (15.43%), civil servants (14.09%), and health workers (8.05%), which is more in this study (Azlan et al., 2020; L. Wang et al., 2020). Consistent with the previous study conducted in Wuhan (D. Wang et al., 2020), this present study also explored males who are mainly visited in the workplace (82.97%, *p* < 0.001) and local market (58.08%, *p* < 0.001). Conversely, males need to attend their daily work for livelihood and go to the local market for their daily needs. Besides, both males (12.66%) and females (16.49%) are early visited in the health care center. Noticeably, no patients had travelled to any other country within the nearest time before getting infected, and only 15% of cases reported the known contact with confirmed cases. Of the confirmed cases, the other frequently observed comorbidities were hypertension (male vs. female: 7.29% vs. 2.06%, *p* = 0.060), diabetes mellitus (male vs. female: 4.86% vs. 6.18%, *p* = 0.612), asthma (male vs. female: 8.33% vs. 5.15%, *p* = 0.306), cancer (male vs. female: 0.01% vs. 1.00%, *p* = 0.745) and pregnancy (female: 5.26%, *p* < 0.001). However, they were observed with a lower percentage than the preceding observation in China and Italy (Colaneri et al., 2020; R. Huang et al., 2020; H. Meng et al., 2020). Maximum patients take necessary vaccines such as Polio (87.9%), BCG (86.2%), Tetanus (82.5%), Mumps (54%) were taken in life. About 37.73% of cases were asymptomatic confirmed COVID-19 cases in the southeast region (*p* = 0.001). Only 5% of asymptomatic cases were observed in Beijing (Tian et al., 2020), and this asymptomatic characteristic was more prevalent among females (40.51%) than that of male (36.84%) patients (*p* = 0.001), which is similar to our findings. We recorded all the identified signs and symptoms of the symptomatic cases. Out of these, the most dominant symptom was cough (34.05%), followed by fever (29.45%). Other symptoms of fatigue (21.78%), headache (13.50%), sore throat (8.28%), ache (12.58%), and diarrhea (3.07%) were also found among patients. In the meanwhile, several studies conducted in Wuhan (Y. Meng et al., 2020), Shanghai (Yu et al., 2020), Beijing (Niu et al., 2020), Hubei (Yang et al., 2020), Jiangsu (R. Huang et al., 2020), and United States (Aggarwal et al., 2020) found that fever and cough as dominant clinical symptoms. However, 29.45% of our study’s symptomatic patients showed comparatively higher fever than that in South Korea (11.6%) (Kim et al., 2020).

There have been no published studies on sex-specific demographics, clinical characteristics, and comorbidity of COVID-19 infected patients, Bangladesh. This work is the first cross-sectional study and the most comprehensive assessment and robust evidence of patients clinical characteristics and comorbidities with COVID-19. Albeit this is a novel study, several limitations might be noted in the present study. Firstly, the present study was performed only in a single institution by obtaining data when patients came for coronavirus testing. Hence, the represented data does not give the whole scenario of all COVID-19 patients of the country. Secondly, the limited number of study populations, especially for female patients. Finally, we did not include any data from hospitalized patients and laboratory outputs. Further investigations are required by the use of enormous raw data and secondary COVID-19 data in the study. However, our study analyzed the demographic and clinical characteristics of COVID-19 patents that aid in identifying possible risk factors and reducing the risk of COVID-19 susceptibility to control this outbreak.

## 5 Conclusions

Nowadays, COVID-19 is a contagious disease and led to a major global health concern. Due to the dynamic nature of the pandemic, more and continuous epidemiological models are needed for forecasting and preparedness in different scenarios. In the present study, we found that males were more affected than female patients, and middle-aged people were mainly affected in both sexes. Fever, cough, and tiredness were the common symptoms found among patients, and several comorbidities (diabetes, hypertension, and asthma) were present among patients.

Maximum COVID-19 patents were B (+ve), and businessmen were mostly affected by this infectious virus. We also observed that the coronavirus mainly affected male patients due to the earlier visit to the workplace, local market, and bank. As the epidemic situation in Bangladesh worsens by the day, reinforcing all of the country’s mitigation measures and laws, as well as adhering to all WHO guidelines, is critical at this time to maintain the exposure probability and transmission rate as low as possible. We anticipate this study will raise the knowledge about COVID-19 and help to take effective and proper decisions to identify risk factors to control this pandemic.

## Data Availability

There were various sections in the survey including demographic information, clinical characteristics, pre-existing medical condition, blood group, earlier visited information, vaccination, etc. The investigators were told that their participation was anonymous and entirely voluntary, and there was no reward for taking part. All the participants willingly participated in this work by giving written permission. They were invited to complete the questionnaire. We were present on hand to answer questions or clarify any doubts that they might have. All filled questionnaires were collected by us one by one.

## Funding disclosure

This work was supported by the Research Cell, Noakhali Science and Technology University, Bangladesh.

## Ethical approval

The study protocol was approved by the institutional ethical clearance committee of Noakhali Science and Technology University, Bangladesh (Approval ID: 44-2020).

## Acknowledgments

We acknowledge the sincere help extended by several COVID-19 Diagnostic Lab Volunteers to name a few are: Md. Sahedul Islam, Md. Amzad Hossain, Mahmudul Islam Rakib, Golam Shamdani, Amor Chandra Nath, and Md. Julker Nyne, of Noakhali Science and Technology University, Bangladesh.

## Notes

### Competing Interest Statement

The authors have declared no competing interest.

### Summary of Updates

Author affiliations updated

